# Peginterferon-lambda for the treatment of COVID-19 in outpatients

**DOI:** 10.1101/2020.11.09.20228098

**Authors:** Jordan J. Feld, Christopher Kandel, Mia J. Biondi, Robert A. Kozak, Muhammad Atif Zahoor, Camille Lemieux, Sergio M. Borgia, Andrea K. Boggild, Jeff Powis, Janine McCready, Darrell H. S. Tan, Tiffany Chan, Bryan Coburn, Deepali Kumar, Atul Humar, Adrienne Chan, Braden O’Neil, Seham Noureldin, Joshua Booth, Rachel Hong, David Smookler, Wesam Aleyadeh, Anjali Patel, Bethany Barber, Julia Casey, Ryan Hiebert, Henna Mistry, Ingrid Choong, Colin Hislop, Deanna M. Santer, D. Lorne Tyrrell, Jeffrey S. Glenn, Adam J. Gehring, Harry L.A. Janssen, Bettina Hansen

**Author notes:** **Correspondence:** Jordan J. Feld MD MPH, Toronto Centre for Liver Disease, Toronto General Hospital, University Health Network, 200 Elizabeth Street, 9EB 240, Toronto, ON, M5G 2C4. Contributed equally to the work (Jordan J. Feld, Christopher Kandel, Mia J. Biondi). ClinicalTrials.gov Identifier: NCT04354259. **Disclosures:** JJF reports research support unrelated to this work from Eiger BioPharmaceuticals. BC has received research support unrelated to this work from Nubiyota LLC and Sanofi. IC and CH are employees of Eiger BioPharmaceuticals. JSG is a board member and founder of Eiger BioPharmaceuticals, Inc., in which he has an equity interest, and is an inventor on a patent application for the use of interferon lambda to treat coronavirus infections. **Author Contributions: Study Concept and Design:** JJF, CK, MJB, DHST, AH, BC, JSG, BH. **Acquisition of Data:** JJF, CK, MJB, RAK, CL, SMB, AKB, JP, JM, DHST, TC, DK, AC, BO, SN, JB, RH, DS, AP, WA, BB, DJ, DS, JC, RH, HM, AZ. **Drafting of the Manuscript:** JJF, CK, MJB, RAK, BH, HLAJ. **Critical Revision of the Manuscript:** JJF, CK, MJB, RAK, SMB, AKB, JP, DHST, BC, BO, WA, IC, CH, DMS, DLT, JSG, AJG, HLAJ, BH. **Statistical Analysis:** JJF, CK, MJB, HLAJ, BH. **Study Supervision:** JJF, CK, MJB, RAK, SN, JB, RH, DS, WA, AP, BB, DJ, DS.

## Abstract

**Background:** There are currently no effective treatments for outpatients with coronavirus disease 2019 (COVID-19). Interferon-lambda-1 is a Type III interferon involved in the innate antiviral response with activity against respiratory pathogens.

**Methods:** In this double-blind, placebo-controlled trial, outpatients with laboratory-confirmed COVID-19 were randomized to a single subcutaneous injection of peginterferon-lambda 180μg or placebo within 7 days of symptom onset or first positive swab if asymptomatic. The primary endpoint was proportion negative for SARS-CoV-2 RNA on Day 7 post-injection.

**Findings:** There were 30 patients per arm, with median baseline SARS-CoV-2 viral load of 6.71 (IQR 1.3-8.0) log copies/mL. The decline in SARS-CoV-2 RNA was greater in those treated with peginterferon-lambda than placebo (p=0.04). On Day 7, 24 participants (80%) in the peginterferon-lambda group had an undetectable viral load compared to 19 (63%) in the placebo arm (p=0.15). After controlling for baseline viral load, peginterferon lambda treatment resulted in a 4.12-fold (95CI 1.15-16.7, p=0.029) higher likelihood of viral clearance by Day 7. Of those with baseline viral load above 10E6 copies/mL, 15/19 (79%) in the peginterferon-lambda group were undetectable on Day 7 compared to 6/16 (38%) in the placebo group (p=0.012). Adverse events were similar between groups with only mild reversible transaminase elevations more frequently observed in the peginterferon-lambda group.

**Interpretation:** Peginterferon-lambda accelerated viral decline in outpatients with COVID-19 resulting in a greater proportion with viral clearance by Day 7, particularly in those with high baseline viral load. Peginterferon-lambda may have potential to prevent clinical deterioration and shorten duration of viral shedding.

(NCT04354259)

**Funding:** This study was supported by the Toronto COVID-19 Action Initiative, University of Toronto and the Ontario First COVID-19 Rapid Research Fund. Medication was supplied by Eiger BioPharma.

**Research in Context:** Treatment trials for COVID-19 have largely focused on hospitalized patients and no treatments are approved for people with mild to moderate disease in the outpatient setting. A number of studies in ambulatory populations have been registered but no controlled studies in the outpatient setting have been reported to date (Pubmed Search October 20, 2020, COVID-19 treatment; controlled trials). Uncontrolled case series of hydroxychloroquine with or without azithromycin have been reported with mixed results but no clear signal of efficacy and some concerns raised about cardiac toxicity. Treamtent in the outpatient setting has potential to prevent infected individuals from deteriorating and perhaps more importantly, may shorten the duration of viral shedding, reducing the risk of transmission and the duration required for self-isolation, with significant public health and societal impact.

**Added value of this study:** This is the first study to show an antiviral effect in outpatients with COVID-19. After controlling for baseline viral load, those treated with peginterferon-lambda had a 4.12-fold (95%CI 1.15-16.7, p=0.029) higher odds of viral clearance by Day 7 compared to those who received placebo. The viral load decline was faster with pegterferon-lambda and the effect was most pronounced in those with high viral loads. In individuals with a baseline viral load of 10E6 copies/mL or higher, 15/19 (79%) in the peginterferon-lambda arm cleared by Day 7 compared to 6/16 (38%) (p=0.012) in the placebo arm (OR 6.25, 95%CI 1.49-31.1, p=0.012), translating to a median time to viral clearance of 7 days (95%CI 6.2-7.8 days) with peginterferon-lambda compared to 10 days (95%CI 7.8-12.2 days) with placebo (p=0.038). Those with low viral loads (<10E6 copies/mL) cleared quickly in both groups. Peginterferon-lambda was well-tolerated with a similar side effect profile to placebo and no concerning laboratory adverse events.

**Implications of all available evidence:** There is no currently approved therapy for outpatients with COVID-19. This study showed that peginterferon-lambda accelerated viral clearance, particularly in those with high baseline viral loads, highlighting the importance of quantitative viral load testing in the evaluation of antiviral agents for COVID-19. Treatment early in the course of disease may prevent clinical deterioration and shorenting of the duration of viral shedding may have important public health impact by limiting transmission and reducing the duration required for self-isolation. Additional trials of peginterferon-lambda and other antiviral strategies in the outpatient setting are required.

## INTRODUCTION

Severe acute respiratory syndrome coronavirus-2 (SARS-CoV-2), the virus causing coronavirus disease 2019 (COVID-19), continues to be a global health threat. To date, only remdesivir and dexamethasone have demonstrated efficacy in randomized trials, but evaluated hospitalized patients only^1, 2^. As with other acute viral infections, early initiation of antiviral therapy for COVID-19 may improve clinical outcomes^3^; yet there are few studies among outpatients. In addition to halting clinical progression, early treatment may shorten the duration of viral shedding, potentially reducing transmission and the duration of self-isolation^4^.

Interferons are produced as part of the innate immune response to viral infections, driving induction of genes with antiviral, antiproliferative and immunoregulatory properties^5^. The broad array of genes induced by interferons limits the risk of antiviral resistance and makes them optimal agents for novel viral pathogens^6^. Interferon-lambdas, known as Type III interferons, exert a similar antiviral state to interferon-alpha/beta, but use a distinct receptor complex with high expression levels limited to epithelial cells in the lung, liver and intestine, resulting in fewer systemic side effects^6^. Interferon-lambda-1 controls respiratory viral infections in mice without the risk of promoting cytokine storm syndrome, as has been seen with Type I interferon treatment^7^. Additionally, interferon-lambda inhibits SARS-CoV-2 replication in cell culture and mouse models^8, 9^.

Peginterferon-lambda, a long-acting form of interferon lambda-1, has been evaluated in over 3,000 patients with viral hepatitis infections, demonstrating similar antiviral efficacy to interferon-alpha, but with an improved side effect profile^10, 11^. We performed a randomized, double-blind, multicenter, placebo-controlled study evaluating treatment with a single 180 μg subcutaneous injection of peginterferon-lambda or placebo in outpatients with COVID-19.

## METHODS

### Study Design

This was a randomized, double-blind, placebo-controlled study, involving six institutions in Toronto, Canada, evaluating treatment with a single 180 μg subcutaneous injection of peginterferon-lambda or placebo in outpatients with COVID-19. The Research Ethics Boards of all participating institutions approved the study, which was registered (NCT04354259) and conducted under a Clinical Trial Application approved by Health Canada. For the complete study protocol and statistical analysis plan, see Supplementary material.

### Participants

Individuals were recruited from outpatient testing centers at six institutions in Toronto, Canada between May 18, 2020 and September 4, 2020. Individuals with SARS-CoV-2-infection confirmed by nasopharyngeal swab were eligible if they were within 7 days of symptom onset or first positive test if asymptomatic. The main exclusion criteria included pregnancy and pre-existing immunosuppressive or other medical conditions that could be worsened by peginterferon-lambda (see protocol for complete inclusion and exclusion criteria). In line with institutional COVID-19 restrictions, all individuals provided witnessed verbal informed consent.

### Randomization and Masking

Eligibile consenting adults were randomized 1:1 to a single subcutaneous injection of 180 μg of peginterferon-lambda or saline placebo. A computer-generated randomization list was created by the study statistician (BH) with a randomization schedule in blocks of four. At the time of randomization, the study personnel received a sealed opaque envelope with the treatment allocation number that indicated which vial to administer to the participant. Numbered study medications were stored in indvidual opaque bags in the study refrigerstor until use. Because of the lack of an identical matching placebo, one of two study personnel administering the medication was aware of the treatment allocation. The participant was instructed to look away during the administration and there were no identifiable features on the syringe to unmask allocation to the participant. After administering the medication, all further follow-up (phone calls and study visits) was done by other study personnel unaware of treatment allocation. A second copy of sealed envelopes with treatment allocation was stored in a locked cabinet for emergency purposes in case of the need for unmasking. Aside from the nurse administering the study medication, all other study personnel and study participants remained masked to treatment allocation until unblinding of the study. The study statistician maintained the masked randomization list on a secure server. Unmasked data were provided to the DSMC for scheduled safety review. No interim efficacy review was performed by the DSMC. Analysis of study results was performed after the statistical analysis plan was finalized, at which point the randomization list linking data to study identification numbers was unmasked.

### Procedures

Study participants attended an outpatient clinic where they were randomized using a computer-generated randomization schedule in blocks of 4. Prior to receiving the study treatment, participants were instructed on proper technique for self-collection of mid-turbinate nasal swabs and collected an observed swab. Treatment with peginterferon-lambda or placebo was administered by a study nurse and participants were observed for 30-60 minutes afterwards. Participants were given written and video instructions to reinforce technique in swab self-collection and whenever possible, swab collection was observed virtually by study staff. Temperature monitoring and assessments of adverse events (AEs) were conducted remotely by study personnel blinded to study arm along with self-collected mid-turbinate (MT) swabs on Day 0, 1, 2, 3, 5, 7, 10 and 14. Hematological and biochemical tests were obtained on Days 0, 3, 7 and 14. Quantitative SARS-CoV-2 results were generated for MT swabs (see Supplementary material). Anti-SARS-CoV-2 IgG antibodies to S protein were measured on Day 0, 3, 7 and 14 (DiaSorin, Saluggia, Italy).

### Outcomes

The primary efficacy outcome was the proportion of individuals with a negative MT swab for SARS-CoV-2 at Day 7, which was chosen based on available data of clearance rates by Day 7 with interferon-beta and a pragmatic consideration that clearance beyond 7 days in an outpatient population would be of limited practical benefit^12^. The primary safety outcome was the incidence of treatment-emergent severe adverse events (SAEs) by Day 14. Secondary outcomes included: time to undetectable SARS-CoV-2 RNA, quantitative change in SARS-CoV-2 RNA over time, anti-SARS-CoV-2 IgG antibody positivity, the incidence and severity (mild/moderate/severe) of AEs, and the proportion hospitalized by Day 14. Detailed directed and open-ended symptoms were assessed serially by phone. Due to overlap between symptoms of COVID-19 and potential peginterferon-lambda AEs, all symptoms were recorded and categorized, and any symptoms outside of the directed symptom assessment were considered AEs. Laboratory AE severity was graded using the Common Terminology Criteria for Adverse Events Version 5.0. An independent Data and Safety Monitoring Committee (DSMC) reviewed safety data after 10, 20 and 30 patients completed 7 days of post-treatment follow-up. After review, the DSMC advised the study team whether to continue enrolment.

### Statistical Analysis

Based on early COVID-19 studies^12, 13^, we estimated 40% viral clearance by Day 7 in the placebo arm and 80% in the peginterferon-lambda arm, requiring 30 patients per arm, to achieve 80% power with alpha of 0.05, accounting for 10% dropout.

Demographic and baseline clinical characteristics were summarized using means with standard error or medians with interquartile ranges for continuous variables, and proportions for categorical variables. The main efficacy outcome was analyzed by a Chi-square test following an intention-to-treat principle. Prespecified analysis of the primary endpoint adjusted for baseline viral load using bivariate logistic regression was performed. Secondary outcomes were compared using Chi-square tests for proportions or Wilcoxon/linear regression controlling for baseline values. The effect of treatment, baseline factors and viral load on clearance by Day 7, were assessed by logistic regression and time-to-clearance by Kaplan-Meier survival analysis. Generalized estimating equations and generalized linear models were used to analyse differences between peginterferon-lambda and placebo in symptom/AE severity grade, and laboratory and viral load patterns over time, respectively. All statistical analyses were performed using SAS 9.4.

## RESULTS

### Participants

Of 364 individuals approached for the study, 105 did not meet inclusion/exclusion criteria and 199 eligible individuals declined to participate (Figure 1). All 60 randomized individuals received an injection and 59 (98%) completed follow-up, with one lost to follow-up after Day 3. The median age was 46 years (IQR 32-54), 35 (58%) were male and eleven (19%) participants were asymptomatic. The mean time from symptom onset to randomization was 4.5±1.7 days. The median baseline SARS-CoV-2 RNA level was 6.71 (IQR 1.3-8.0) log copies/mL with 10 (33%) individuals in the placebo group and 5 (17%) in the peginterferon-lambda group having undetectable viral load on the day of randomization. Other baseline characteristics were similar between groups (Table 1).

**Table 1.** Baseline Characteristics in the peginterferon-lambda and placebo groups.

|  | **Peginterferon-Lambda**<br>**n=30** | **Placebo**<br>**n=30** |
| --- | --- | --- |
| Female, n(%) | 18 (60) | 17 (57) |
| Age [years], median (IQR) | 48 (30-53) | 39 (33-55) |
| Race/Ethnicity n(%) |  |  |
| White | 15 (50) | 16 (53) |
| Black | 1 (3) | 5 (17) |
| Asian | 8 (27) | 7 (23) |
| Other | 6 (20) | 2 (7) |
| Co-morbidity* n(%) | 5 (17) | 4 (13) |
| Body Mass Index, kg/m<sup>2</sup> mean(sd) | 27.3 (5.2) | 26.1 (4.2) |
| Body Mass Index Category, n(%) |  |  |
| <25 kg/m<sup>2</sup> | 9 (30) | 11 (37) |
| 25-30 kg/m<sup>2</sup> | 15 (50) | 13 (43) |
| >30 kg/m<sup>2</sup> | 6 (20) | 6 (20) |
| Interferon lambda 4 genotype |  |  |
| TT | 18 (60) | 16 (57) |
| Non-TT | 12 (40) | 12 (43) |
| Asymptomatic | 5 (17) | 6 (20) |
| Symptom onset to injection (days), mean(sd) | 4.3 (1.7) | 4.7 (1.7) |
| Positive SARS-CoV-2 test to injection (days), mean(sd) | 3.2 (1.1) | 3.3 (1.2) |
| Baseline laboratory results |  |  |
| Hemoglobin (g/L), mean(sd) | 14.7 (1.4) | 14.9(1.6) |
| White Blood Cells (x10e9/L ), mean(sd) | 4.9 (2.1) | 5.1 (1.7) |
| Lymphocytes (x10e9/L), mean(sd) | 1.5 (0.4) | 1.5 (0.5) |
| Neutrophils (x10e9/L), mean(sd) | 2.9 (1.8) | 3.1 (1.6) |
| Platelets (x10e9/L), mean(sd) | 221 (62) | 213 (64) |
| Creatinine (μmol/L), mean(sd) | 80 (14) | 81 (18) |
| Alanine aminotransferase (U/L), mean(sd) | 32(16) | 39 (52) |
| Aspartate aminotransferase (U/L), mean(sd) | 28 (11) | 32 (24) |
| Total bilirubin (μmol/L), mean(sd) | 10 (5) | 12 (10) |
| Baseline SARS-CoV-2 viral load (log copies/mL), mean(sd) | 6.16 (3.14) | 4.87 (3.68) |
| SARS-CoV-2 RNA undetectable at baseline, n(%) | 5 (17) | 10 (33) |
| SARS-CoV-2 RNA ≥ 10E6 copies/mL at baseline, n(%) | 19 (63) | 16 (53) |
| anti-SARS-CoV-2 S protein IgG antibody at baseline, n<sup>#</sup>(%) | 0/27 (0) | 5/24 (21) |

\* Hypertension, diabetes mellitus, chronic obstructive pulmonary disease, heart disease

**Figure 1.**
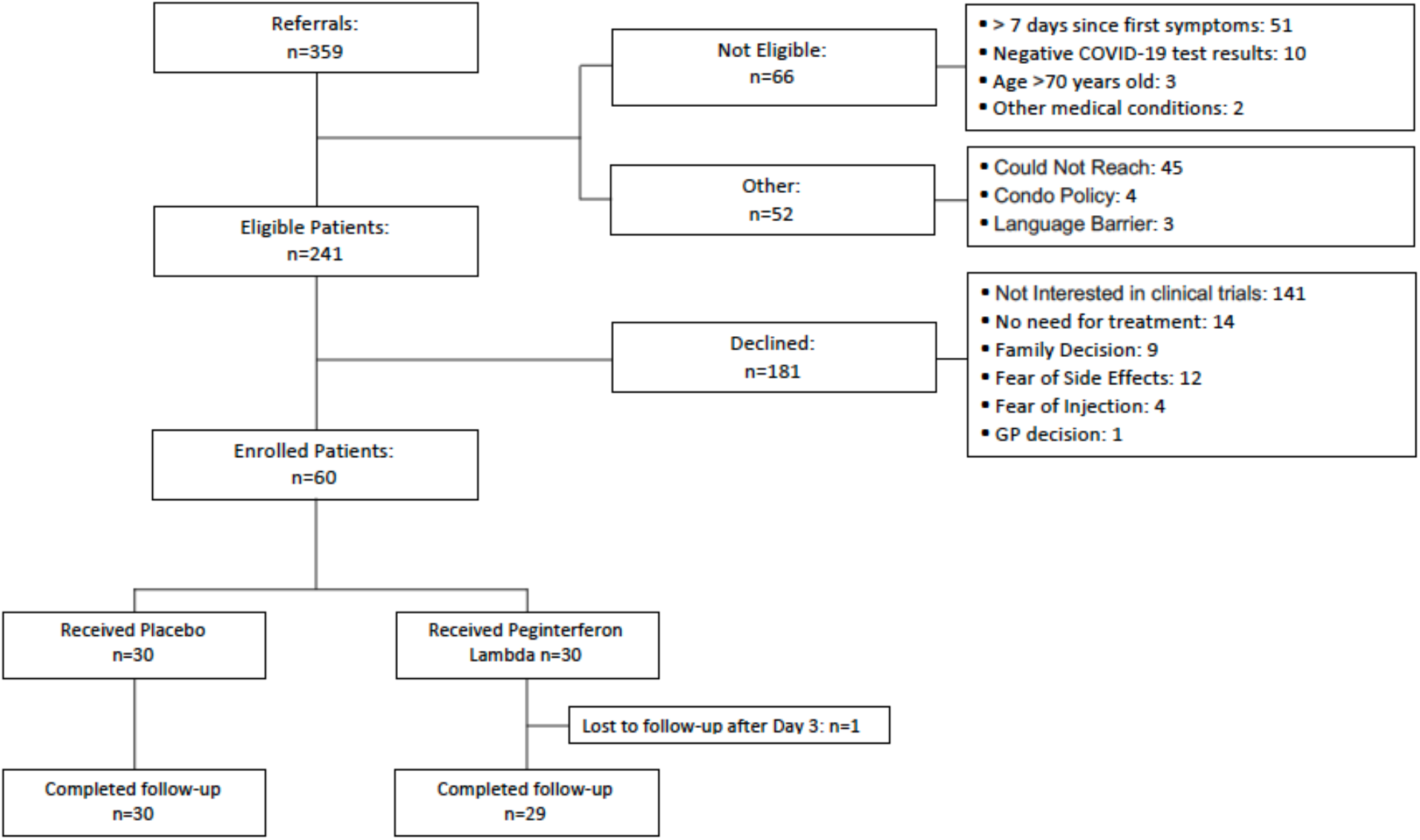
CONSORT diagram. The CONSORT diagram detailing the screening, randomization and study completion.

### Outcomes

The baseline SARS-CoV-2 RNA level was higher in the peginterferon lambda group and was significantly associated with the probability of clearance by Day 7 (OR 0.69, 95%CI 0.51-0.87, p=0.001). Overall, by Day 7, 24/30 (80%) in the peginterferon-lambda group were negative for SARS-CoV-2 RNA compared to 19/30 (63%) in the placebo arm (p=0.15) (Figure 2a). After adjusting for baseline viral load, peginterferon-lambda treatment was significantly associated with clearance by Day 7 (OR=4.12, 95%CI 1.15-16.7, p=0.029) (Table 2).

**Table 2.**
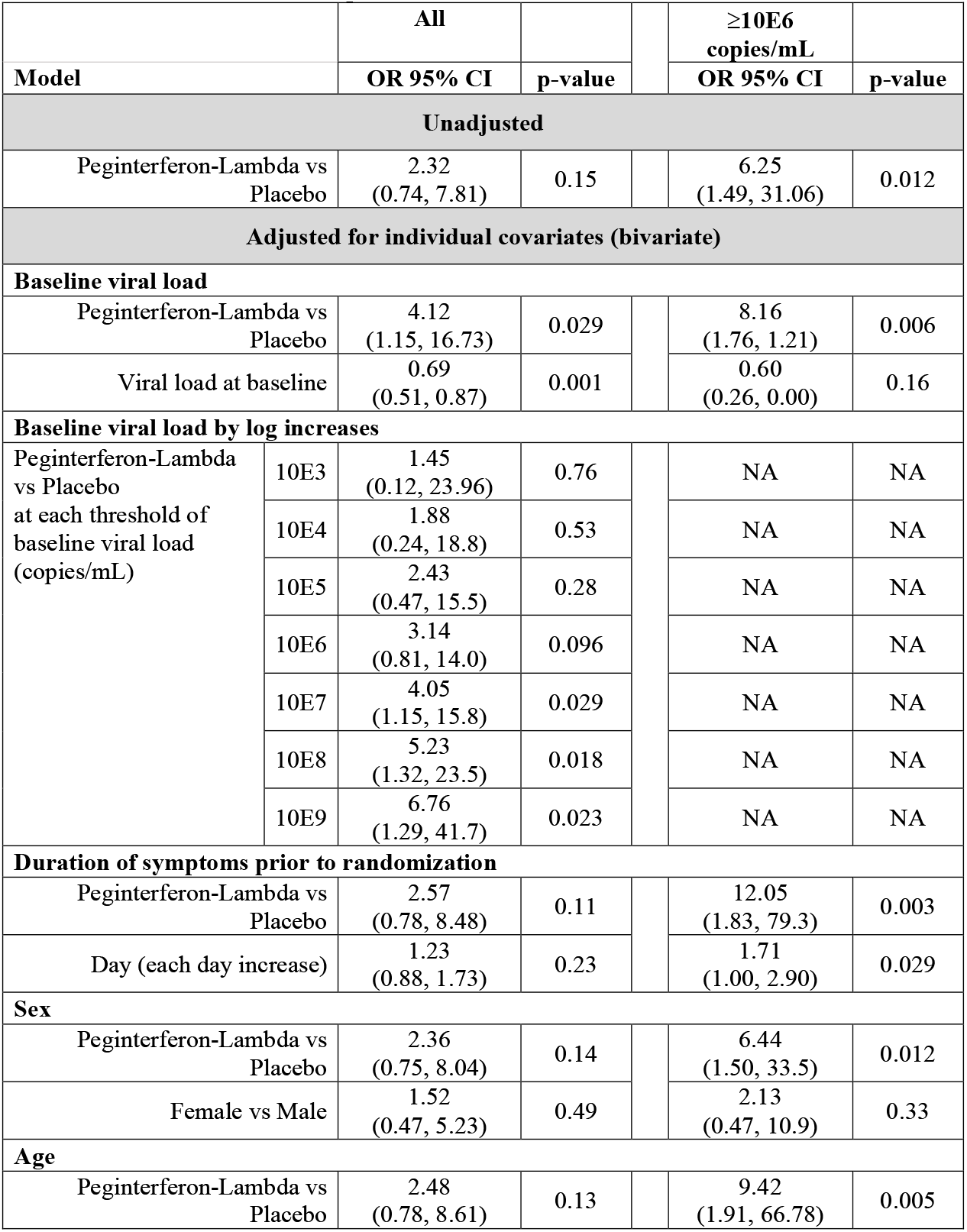
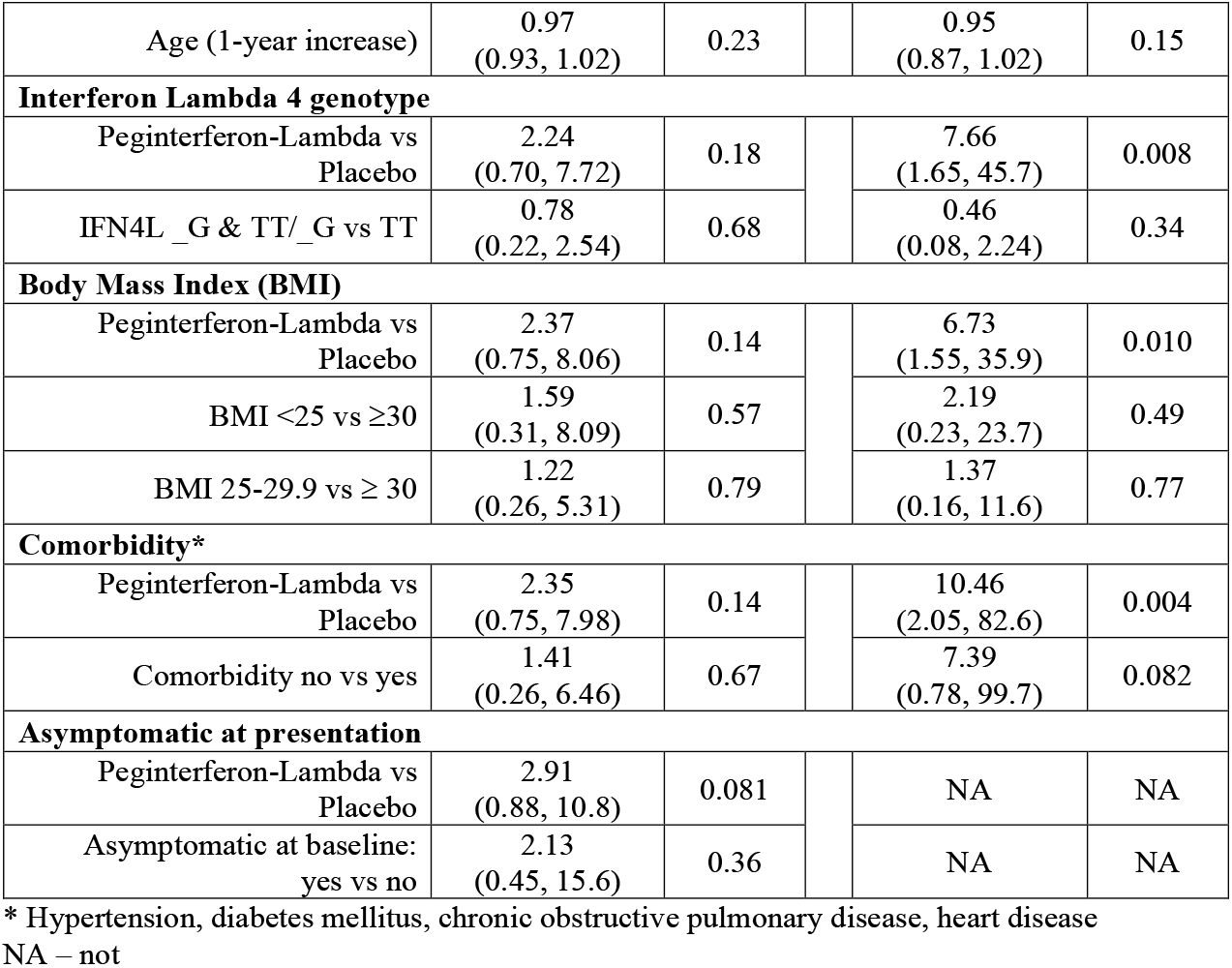
Crude and adjusted odds of undetectable SARS-CoV-2 RNA at Day 7 with peginterferon-lambda compared to placebo treatment in entire cohort and in subgroup with baseline viral load ≥10E6 copies/mL.

**Figure 2.**
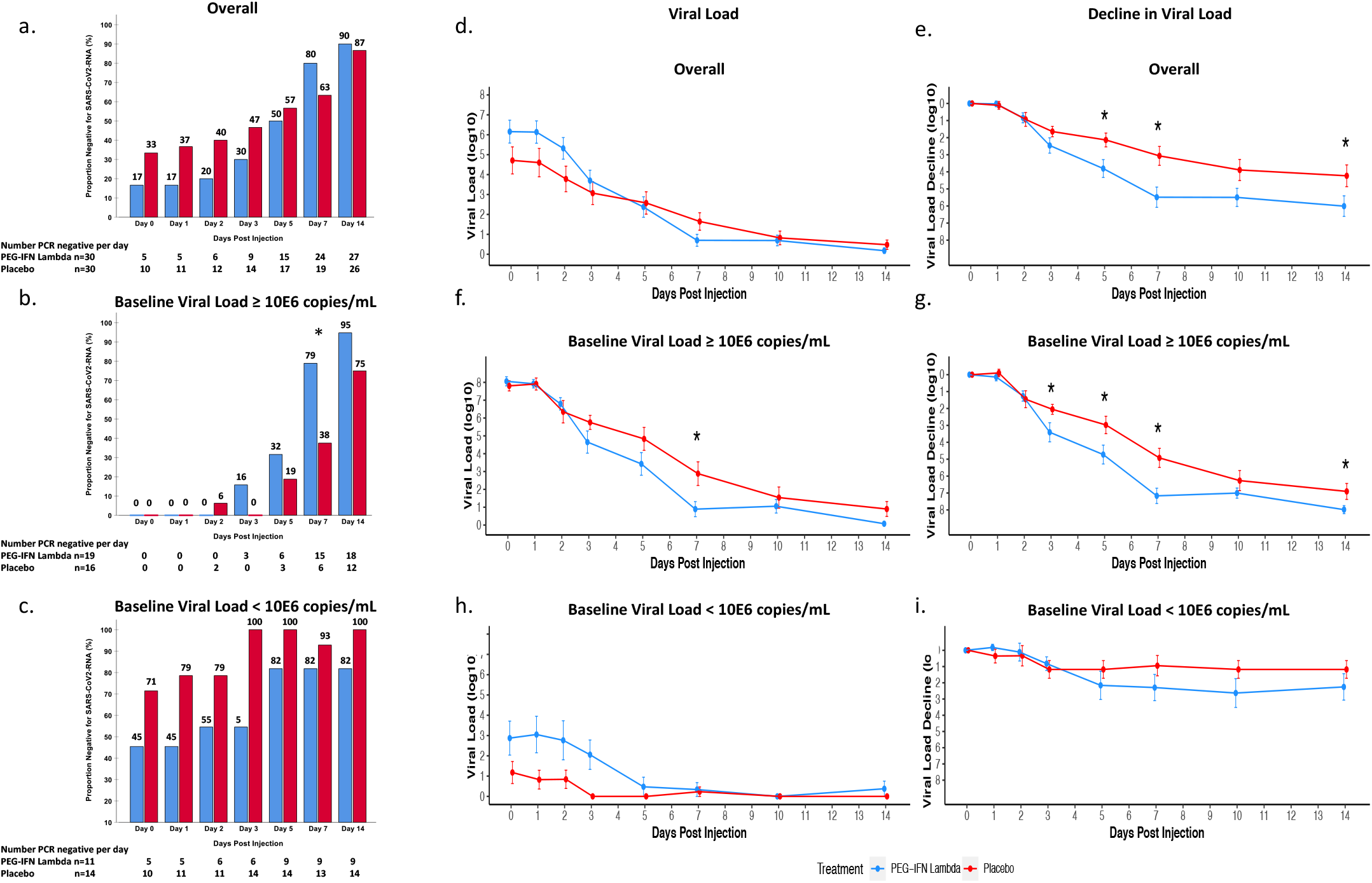
Proportion negative for SARS-CoV-2 RNA over time and mean absolute and change in SARS-CoV-2 viral load over time. The proportion of patients negative for SARS-CoV-2 RNA per day post-injection, including all treated patients (Panel a) and stratified by baseline viral load, above (Panel b) and below 10E6 copies/mL (Panel c). The mean SARS-CoV-2 viral load (Panels d, f, and h) and viral load decline from baseline (Panels e, g, and i) for the peginterferon-lambda and placebo groups per day post-injection and stratified by baseline viral load above or below 10E6 copies/mL. The I bars represent standard error. * p <0.05 at indicated time points. In panel b, p=0.013 at Day 7. In panel e, p=0.04 for trend and at Day 3, p=0.14, Day 5 p=0.013, Day 7 p=0.004, Day 14 p=0.048. In Panel f, p=0.017 at Day 7. In Panel g, difference at Day 3 p=0.042, Day 5 p=0.029, Day 7 p=0.004, Day 14 p=0.039.

The odds of viral clearance by Day 7 with peginterferon-lambda treatment compared to placebo increased with every log increase in baseline viral load (Figure 3). For those with baseline RNA of 10E6 copies/mL or greater, the proportion undetectable at Day 7 in the peginterferon-lambda group was 15/19 (79%) compared to 6/16 (38%) in the placebo group (OR 6.25, 95%CI 1.49-31.1, p=0.012) (Figure 2b), translating to a median time to viral clearance of 7 (95%CI 6.2-7.8) days with peginterferon-lambda compared to 10 (95%CI 7.8-12.2) days with placebo (p=0.038) (Supplementary Figure 1). Of those still positive at Day 7, participants in the peginterferon lambda group had lower viral loads than those in the placebo group, with 3 of 4 at 10E4 copeis per mL or lower, compared to 6/10 (60%) above 10E5 copies per mL in the placebo group (Supplementary Table 2). In contrast, in those with baseline viral load below 10E6 copies/mL, viral loads were higher in the peginterferon-lambda group, but both groups cleared very quickly; 9/11 (82%) in the peginterferon-lambda arm and 13/14 (93%) in the placebo arm were undetectable at Day 7 (OR 0.35, 95%CI 0.01-4.15, p=0.40) (Figure 2c).

**Figure 3.**
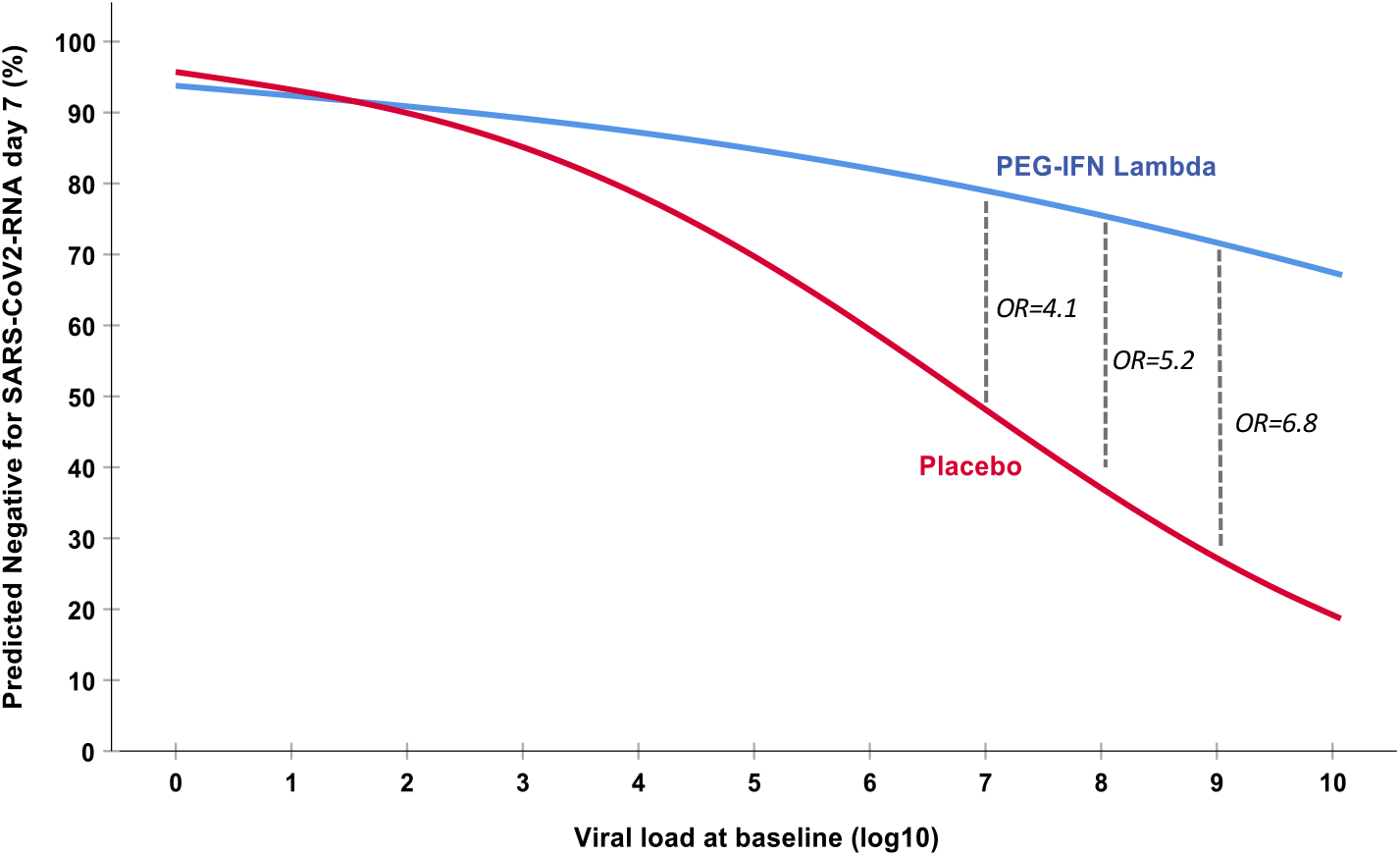
Probability of SARS-CoV-2 clearance by Day 7 according to baseline viral load. The odds of clearance by Day 7 in the peginterferon-lambda group as compared to the placebo group for each baseline viral load (log copies/mL).

The mean decline in SARS-CoV-2 RNA was significantly greater in the peginterferon-lambda than in the placebo group from Day 5 onwards (Figure 2e), with a similar effect observed when restricted to those with detectable virus at baseline (Supplementary Figure 2) or baseline viral load at or above 10E6 copies/mL (Figure 2g). Clearance was rapid in the low viral load group in both groups (Figure 2i).

No baseline covariates modified the association between baseline viral load and treatment assignment with clearance by Day 7 (Table 2, Supplementary Figure 3). Participants who were asymptomatic were more likely to have baseline viral loads below 10E6 copies/mL than those with symptoms (91% vs 27%, p<0.001). At randomization, 5/51 (9.7%) participants with available samples were seropositive for SARS-CoV-2 S IgG antibodies, of whom 4 had undetectable SARS-CoV-2 RNA. Antibody positivity increased in both groups over time (Supplementary Figure 4). The presence of antibodies at any timepoint was associated with a corresponding lower viral load, with the association weakening with time as people cleared.

**Figure 4.**
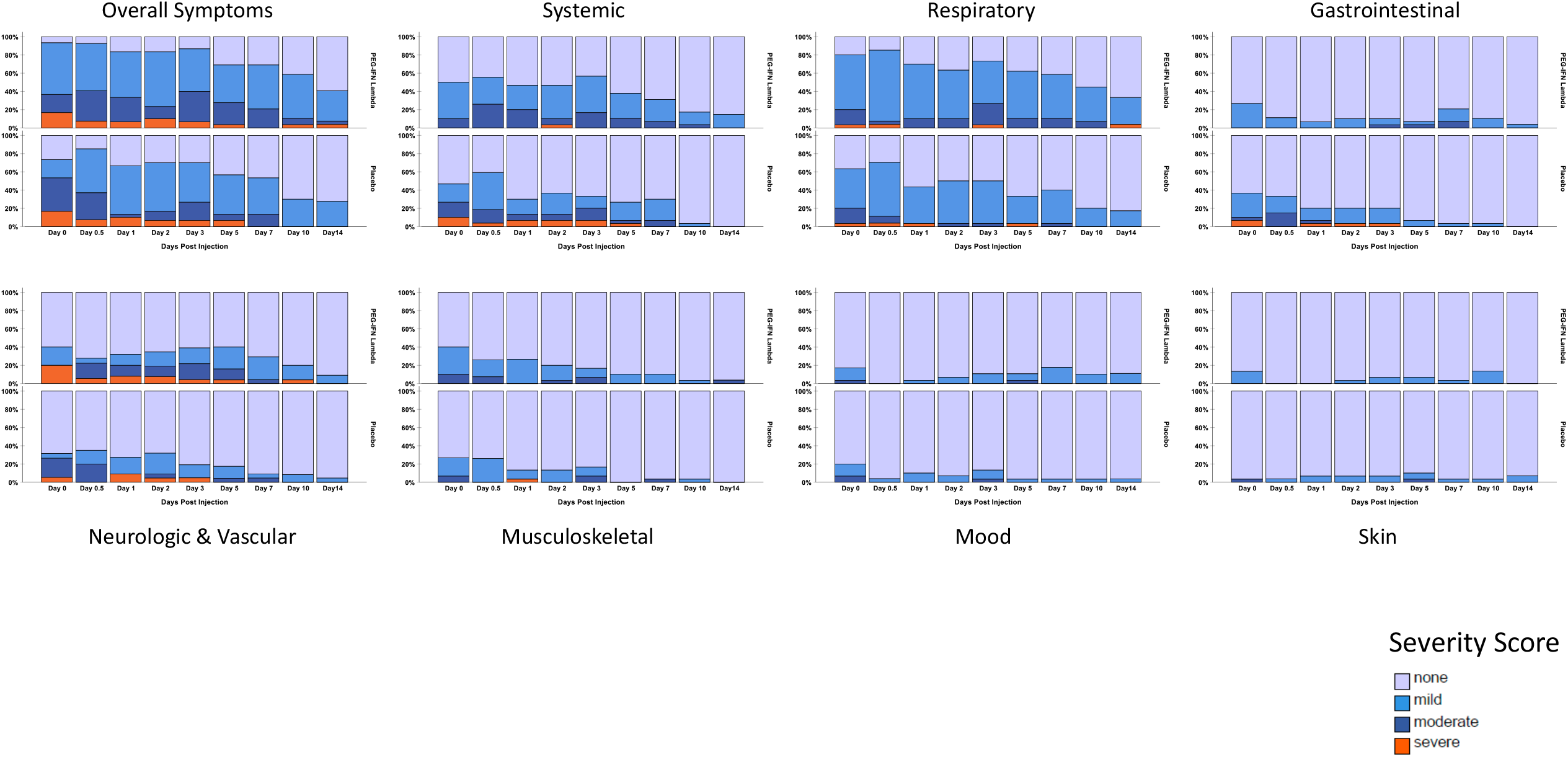
Symptoms in the peginterferon-lambda and placebo groups per day. The proportion of participants reporting no (light blue), mild (blue), moderate (dark blue) or severe symptoms (orange) is shown for the peginterferon-lambda group and the placebo group. Symptoms were grouped into categories and the most severe ranking of any symptom in the specific category was used for each participant at each day. Overall significant declines of symptom severity over time were observed in all categories in both groups (p<0.0001), except Skin. No significant difference between treatments and no significant difference of decline of symptom severity between treatments were observed.

### Safety

Symptoms were grouped into 7 categories (Table 3) and reported as absent/mild/moderate or severe. Respiratory and fever/systemic symptoms were most common in both groups (Figure 4). Documented temperature above 38°C was rare but only reported beyond Day 2 in the peginterferon-lambda group (Supplementary Figure 5). Overall, most symptoms in both groups were mild and there was no difference in frequency, severity or rate of improvement of any of the 7 symptom categories between treatment groups. (Figure 3, Supplementary Table 2). A symptom was graded as severe on 20 occasions by 7 patients in the peginterferon-lambda group and on 30 occasions by 7 patients in the placebo group with somewhat different patterns. In the peginterferon group, severe symptoms were most commonly loss of taste/smell whereas in the placebo group, fever/systemic symptoms were most frequently rated as severe (Supplementarry Table 3). Symptoms improved in both groups over time at a similar rate (Supplementary Table 2, Figure 3). Participants with baseline viral loads above 10E6 copies/mL had higher symptom scores than those with low baseline viral loads in all categories, except skin symptoms, but improved similarly during follow-up (Supplementary Table 2).

**Table 3.** Categorization of symptoms that were assessed daily into whether they were likely attributed to COVID-19, peginterferon-lambda or either/both.

| Symptom Category | Symptom | COVID-19 | Peginterferon-Lambda |
| --- | --- | --- | --- |
| **Systemic** | Fever | X | X |
|  | Chills | X | X |
|  | Rigors | X | X |
|  | Fatigue | X | X |
| **Respiratory** | Cough | X |  |
|  | Sore Throat | X |  |
|  | Shortness of breath | X |  |
|  | Chest pain | X |  |
|  | Runny nose | X |  |
|  | Conjunctivitis | X |  |
| **Gastrointestinal** | Abdominal pain | X |  |
|  | Nausea | X |  |
|  | Vomiting | X |  |
|  | Diarrhea | X |  |
| **Musculoskeletal** | Muscle Pain | X | X |
| **Skin** | Rash |  | X |
|  | Itch |  | X |
|  | Injection Site Reaction |  | X |
| **Mood** | Depressed Mood | X | X |
| **Neurologic and Vascular** | Loss of smell | X |  |
|  | Loss of taste | X |  |
|  | Change in color of fingers/toes | X |  |

**Figure 5.**
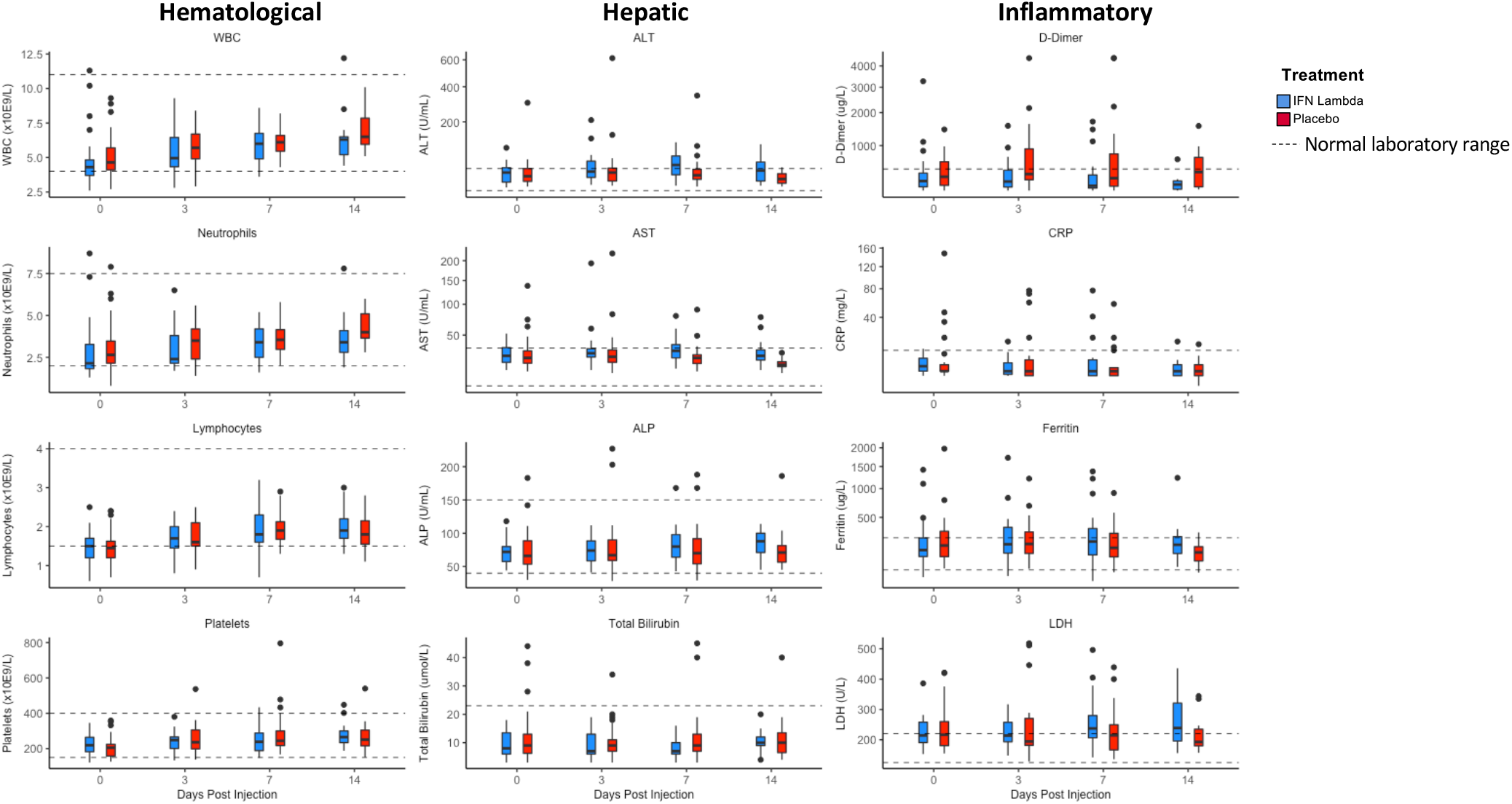
Laboratory values in the peginterferon-lambda and placebo groups per day. The median and interquartile ranges for hematological, hepatic and inflammatory markers at Day 0, 3, 7 and 14 in each group.

Laboratory AEs were mild and similar between groups. Aminotransferases were elevated at baseline in 3 (11%) participants in both groups and increased mildly, moreso in the peginterferon-lambda group. However, only two individuals met the threshold of Grade 3 elevation, one in each arm. No other grade 3/4 laboratory AEs were reported (Table 4). There were no elevations in bilirubin with the observed increases in aminotransferases. Hemoglobin, white blood count and platelets were similar with no episodes of myelosuppression in either group. D-dimers were elevated in both groups at baseline but declined over time only in the peginterferon-lambda group (Day 7: placebo 841 ug/L vs peginterferon-lambda 437 ug/L, p=0.02). Other inflammatory markers including ferritin and C-reactive protein were elevated at baseline in both groups and changed minimally over time (Figure 5).

**Table 4.** Summary of the Adverse Events (AEs) and Severe Adverse Events (SAEs) by treatment group.

| Type of Event | Intervention |  |
| --- | --- | --- |
|  | Peginterferon-Lambda | Placebo |
| **Severe Symptoms** |  |  |
| Reports (number of participants*) | 20 (7) | 30 (7) |
| AEs | 2 | 1 |
| SAEs | 1 | 1 |
| Treatment-Related AEs | 0 | 0 |
| Treatment-Related SAEs | 0 | 0 |
| **Laboratory Abnormalities (grade 3 or 4)** |  |  |
| Hemoglobin | 0 | 0 |
| White Blood Cells | 0 | 0 |
| Lymphocytes | 0 | 0 |
| Neutrophils | 0 | 1 |
| Platelets | 0 | 0 |
| Creatinine | 0 | 0 |
| Alanine aminotransferase (ALT) | 1 | 3 |
| Aspartate aminotransferase (AST) | 1 | 1 |
| Total bilirubin | 0 | 0 |

AE – adverse event; SAE – severe adverse event
\* Some participants reported multiple severe symptoms

AEs outside of the directed symptom categories occurred in one participant in the placebo arm (rectal bleeding) and in two who received peginterferon-lambda (confusion, pneumonia); all deemed unrelated to treatment. One serious adverse event was reported in each group. A participant in the placebo group was hospitalized on Day 1 post-injection with progressive dyspnea attributed to worsening COVID-19, that improved over time leading to discharge on Day 3. One participant in the peginterferon-lambda group was admitted to hospital on Day 14 with shortness of breath and found to have a pulmonary embolism necessitating anticoagulation. No deaths occurred in either group.

## DISCUSSION

Treatment with a single dose of peginterferon-lambda accelerated viral load decline and, after controlling for baseline viral load, reduced the time to viral clearance in outpatients with COVID-19. The treatment effect was most apparent in those with high baseline viral loads. Peginterferon-lambda was well tolerated with similar symptoms to those treated with placebo.

Results for SARS-CoV-2 diagnostic testing are routinely reported dichotomously as positive or negative, without viral load quantification. Cycle threshold (Ct) values are sometimes reported, but are only semi-quantitative, and vary by assay and even by run, such that results cannot be reliably compared. Our inclusion of plasmid-derived cDNA standards with every PCR run allowed for quantification of SARS-CoV-2 RNA results and robust direct comparison between samples. Quantification is useful clinically as higher viral levels have been correlated with greater severity of COVID-19^14, 15^ and infectivity^16^. As people clear infection, they may have persistently very low levels of RNA detected at very high Ct values (>33), which are not infectious^17^.

We found that the odds of clearance were greater in all study particpants with peginterferon-lambda than with placebo after controlling for baseline viral load. However, the effect of peginterferon-lambda was most evident when baseline viral loads were above 10E6 copies/mL. While the specific threshold for transmissible virus is unknown, using a standard infectivity assay, Bullard and colleagues reported that at Ct values above 24, corresponding to approximately 10E6-10E7 copies/mL, infectious virus could not be detected^16^. We observed that in individuals with low levels of virus, irrespective of their assigned group, spontaneous clearance occurred rapidly and near-universally by Day 7. This does not indicate lack of effectiveness of peginterferon-lambda at low viral loads, but rather that with such low levels of virus, treatment was not required as clearance was imminent. Indeed, 10 participants in the plaebo group and 5 in the peginterferon arm already had undetectable virus by the day of randomization. In addition, 5 participants, all in the placebo arm, had already developed SARS-CoV-2-specific antibodies by the day of randomization. Similarly, recent evaluation of the REGN-COV2 monoclonal antibody cocktail demonstrated that individuals with the highest baseline viral loads exhibited the largest reduction in SARS-CoV-2 RNA with treatment, while those with detectable SARS-CoV-2 antibodies at baseline (45% of the study population) had low viral loads and did not benefit from therapy^18^. It is likely that all antiviral strategies will be most effective early in infection and maximally beneficial to those with highest viral loads. Indeed early reports of interferon-beta use have not shown a clear benefit in hospitalized patients, however whether this relates to late introduction of therapy or possibly to the pro-inflammatory effects of Type I interferon is unknown. Ideally, antivirials would be given shortly after disease onset as rapid reduction of viral load would likely lower the risk of clinical deterioration, but also may reduce transmission, translating into significant public health benefits.

In the placebo group with high baseline viral load, 10/16 (63%) participants had detectable virus at Day 7, with 6/10 (60%) continuing to exceed 10E5 copies/mL, raising concern of persistent shedding of competent virus. In contrast, only 4/19 (21%) participants who received peginterferon-lambda had detectable virus at Day 7, all with viral loads below 10E6 copies/mL. If these results are confirmed in larger studies, either quantitative testing could be introduced, with the added benefit of predicting those at risk of a severe clinical course, or a qualitative assay, ideally a point-of-care test, could be titrated to achieve an analytical sensitivity of ∼10E6 copies/mL allowing for immediate risk stratification and determination of the need for treatment. Indeed, this could likely already be achieved using currently available rapid antigen tests, with detection sensitivities in the range of 10-50,000 copies/mL, safely below the infectious threshold but avoiding those with extremely low viral loads who are unlikely to require any intervention^19^. Alternatively, given the tolerability of a single dose of peginterferon-lambda it may be reasonable to consider treatment irrespective of viral load, as a simple, universal approach.

Peginterferon-lambda was well tolerated with no identified safety concerns. Side effects of peginterferon-lambda overlap with COVID-19 symptoms, making it difficult to distinguish whether AEs were related to treatment or persistent infectious symptoms. As has been reported previously, symptoms were more prominent in those with higher viral loads. With detailed serial symptom assessment, we found that symptoms improved in both groups over time without obvious differences. Notably, among those who were asymptomatic at baseline, there was no difference in AEs between the treatment and placebo groups. Mild, reversible transaminase elevations were seen more frequently in the peginterferon-lambda group, which have been reported previously^10^. Intriguingly, D-dimer levels fell with peginterferon-lambda treatment, which may be relevant given the association of high levels with more severe disease and increased all-cause mortality^20-22^. The side effect profile and absence of hematological toxicity is consistent with the better tolerability of Type III interferons compared to Type I interferons like alpha/beta^10^. Treatment with interferon-lambda may be particularly attractive given reports that impaired interferon production and the presence of autoantibodies to interferon-alpha are associated with severe COVID-19^23-25^. Additional benefits include the broad activity of interferon lambda against multiple respiratory pathogens, including influenza, its very high barrier to resistance and a long-acting formulation that permits a single subcutaneous injection^6,7^.

Study imitations include the small sample size, although clearance rates in those with high viral loads were consistent with the power calculations. Based on viral load and antibody data at the baseline visit, several participants were likely clearing the infection, an observation reported in other COVID-19 outpatient studies^18^. The benefit of treatment was more pronounced in the group with a high baseline viral load, who were earlier in their course of infection. Early treatment would be optimal, or alternatively introduction of quantitative assays or calibrated qualitative tests for COVD-19 diagnosis and risk stratification could be used to identify those most likely to benefit from therapy. As a Phase II trial, the study was not powered to showed differences in transmission, which are very hard to document, or hospitalization, which would require a larger study enriched for those at high risk of complications. However, as a first step to confirm efficacy, viral clearance is a key relevant endpoint. There were more Black participants in the placebo group, a population with reduced responsiveness to Type I interferon for treatment of viral hepatitis. However, there were similar proportions with the treatment-responsive interferon-lambda genotype (TT), which is strongly associated with response to interferon-alpha for hepatitis C infection and thought to explain most of the differential response to interferon by race^26^. No effect of the interferon-lambda genotype was observed on baseline viral load or response to treatment in the interferon-lambda arm. A high proportion of eligible individuals declined to participate in the study, likely due to the listed AE profile, which reflected weekly injections for a year of treatment for hepatitis B and C infections^10, 11^. Importantly, the enrolled population was diverse, with individuals born in 25 different countries.

In conclusion, this is among the first antiviral therapies to show benefit among outpatients with COVID-19. Peginterferon-lambda accelerated viral clearance, particularly in those with high baseline viral load. This treatment may have potential to avert clinical deterioration, shorten the duration of infectiousness and limit required isolation time, with significant public health and societal impact.

## Data Availability

Data will be available for use through contact with the corresponding author after approval of proposed protocol. Contact

