## SupplementalMaterial for "Peginterferon-lambda for the treatment of COVID-19 in outpatients": Supplementary Appendix FINAL.pdf

This appendix has been provided by the authors to give readers additional information about their work.

**Supplementary Appendix**

Supplemental Methods ……………………………………………….....3
Supplemental Figure 1 ………………………………………………... 4
Supplemental Figure 2 …………………………...……………… 5
Supplemental Figure 3 ……………………..…………..……………… 6
Supplemental Figure 4 ……………………...………………………… 7
Supplemental Figure 5 ………………………...……………………… 8
Supplemental Table 1 ……………………………………………….....9
Supplemental Table 2 ………………………………………………... 10
Supplemental Table 3 ……………………………………………………11

### Supplemental Methods

### SARS-CoV-2 Viral Load Testing

A self-collected mid-turbinate swab was obtained. Samples were inactivated and viral nucleic acid was extracted using the NucliSENS EasyMAG platform (bioMerieux, Marcy-l'Etoile, France), and real-time reverse-transcriptase PCR was performed on the Rotorgene Q (Qiagen, Hilden, Germany) using the primers and probes for the SARS-CoV-2 E gene[24,25]. To generate standard curves, dilutions of a synthetic plasmid containing a segment of the E-gene were used (GenScript, USA). Samples with either no Ct value or a value that was below the level of quantification were counted as negative. For the purposes of calculation, values below the limit of quantification (~20 copies/mL) were given an arbitrary value of 10 copies/mL.

**Figure S1. Time to viral clearance in those with baseline viral load ≥ 10E6 copies/mL.** The time to undetectable SARS-CoV-2 RNA is shown for the peginterferon-lambda and the placebo group for participants with a baseline viral load above 10E6 copies/mL. The curves are compared using the log-rank test and the median time to clearance with 95% CI is shown for each group.

**Figure S2. Proportion negative for SARS-CoV-2 RNA over time and mean absolute and change in SARS-CoV-2 viral load over time in those with detectable SARS-CoV-2 at baseline.**

The proportion of patients negative for SARS-CoV-2 RNA per day post-injection in those with a detectable viral load at baseline (Panel a) with the mean SARS-CoV-2 RNA viral load (Panel b) and log decline from baseline (Panel c) over time in participants with a detectable baseline viral load in the peginterferon-lambda treatment (n=25) and placebo groups (n=20).

*Panel b: Difference at Day 7 p=0.042

*Panel c: Difference at Day 5 p=0.036 and Day 7 p=0.006

**Figure S3. Odds ratio for clearance by Day 7 with peginterferon-lambda vs placebo, controlled for individual baseline covariates, in those with baseline viral load ≥ 10E6 copies/mL**

Forest plot showing odds ratio with 95% CI for the odds of clearance by Day 7 with peginterferon lambda treatment compared to placebo, controlled for each individual (bivariate) covariate.

**Figure S4. Presence of SARS-CoV-2 antibodies over time stratified by baseline viral load.**

The proportion of individuals with positive anti-SARS-CoV-2 S protein IgG antibodies per day post-injection, stratified by baseline viral load above or below 10E6 copies/mL.

**Figure S5. Self-collected oral temperature measurements.**

Self-collected oral temperature measurements on days 0, 2, 3, 4, 5, 6, 7, 10, 12 and 14 separated into <38$^{o}$ Celsius, 38-39$^{o}$ Celsius and above 39$^{o}$ Celsius stratified by group.

**Table S1. Residual SARS-CoV-2 RNA levels at Day 7 post-treatment in those with baseline viral loads above and below 10E6 copies/mL**

| **Baseline Viral Load** | **Residual Baseline Viral Load at Day 7 in Copies/mL** | |
|---|---|---|
| | **Peginterferon-Lambda** | **Placebo** |
| **≥ 10E6 copies/mL** | n=4/19 (21%)<br><br>2.74E3<br>4.59E3<br>7.95E3<br>8.58E5 | n=10/16 (62.5%)<br><br>1.61E1<br>3.10E2<br>5.10E3<br>4.28E4<br>1.46E5<br>1.66E5<br>2.62E5<br>4.17E5<br>6.49E5<br>6.40E6 |
| **< 10E6 copies/mL** | n=1/11 (9%)<br><br>2.41E3 | n=1/14 (7%)<br><br>1.82E3 |

**Table S2. Association between peginterferon-lambda treatment vs placebo with specific symptom categories and with symptom evolution over time. Results shown for all participants and stratified by baseline viral load.**

| Symptom Category | Change in severity (day) | All samples | | Baseline Viral Load ≥10E6 copies/mL | | Baseline Viral Load <10E6 copies/mL | |
|---|---|---|---|---|---|---|---|
| | | OR (95% CI) | p-value | OR (95% CI) | p-value | OR (95% CI) | p-value |
| **All Symptoms** | Peginterferon-lambda | 0.71 (0.61, 0.82) | <.0001 | 0.74 (0.62, 0.88) | 0.0007 | 0.68 (0.53, 0.87) | 0.0024 |
| | Placebo | 0.64 (0.55, 0.75) | <.0001 | 0.60 (0.48, 0.74) | <.0001 | 0.76 (0.60, 0.96) | 0.0187 |
| | *Difference in decline* | | 0.32 | | 0.10 | | 0.46 |
| **Fever/Systemic** | Peginterferon-lambda | 0.72 (0.62, 0.84) | <.0001 | 0.77 (0.67, 0.90) | 0.0008 | 0.64 (0.53, 0.77) | <.0001 |
| | Placebo | 0.60 (0.49, 0.73) | <.0001 | 0.64 (0.52, 0.77) | <.0001 | 0.55 (0.43, 0.69) | <.0001 |
| | *Difference in decline* | | 0.06 | | 0.06 | | 0.24 |
| **Respiratory** | Peginterferon-lambda | 0.64 (0.51, 0.80) | <.0001 | 0.70 (0.55, 0.88) | 0.002 | 0.48 (0.26, 0.91) | 0.024 |
| | Placebo | 0.52 (0.39, 0.69) | <.0001 | 0.58 (0.42, 0.78) | 0.0004 | 0.41 (0.20, 0.83) | 0.013 |
| | *Difference in decline* | | 0.19 | | 0.29 | | 0.61 |
| **Gastrointestinal** | Peginterferon-lambda | 0.92 (0.75, 1.15)* | 0.47 | 0.92 (0.83, 1.01)* | 0.091 | 0.98 (0.74, 1.3)* | 0.90 |
| | Placebo | 0.57 (0.42, 0.77) | 0.0003 | 0.53 (0.41, 0.69) | <.0001 | 0.61 (0.42, 0.9) | 0.013 |
| | *Difference in decline* | | 0.0004 | | 0.0001 | | 0.0001 |
| **Musculoskeletal** | Peginterferon-lambda | 0.54 (0.35, 0.83) | 0.005 | 0.51 (0.33, 0.8) | 0.003 | 0.63 (0.46, 0.85) | 0.003 |
| | Placebo | 0.50 (0.32, 0.77) | 0.002 | 0.48 (0.31, 0.75) | 0.001 | 0.48 (0.20, 1.17) | 0.11 |
| | *Difference in decline* | | 0.70 | | 0.81 | | 0.57 |
| **Skin** | Peginterferon-lambda | 0.89 (0.57, 1.38) | 0.59 | 0.79 (0.37, 1.67) | 0.53 | 0.40 (0.02, 9.51) | 0.37 |
| | Placebo | 0.88 (0.57, 1.35) | 0.56 | 0.50 (0.16, 1.60) | 0.24 | 0.91 (0.73, 1.13) | 0.22 |
| | *Difference in decline* | | 0.96 | | 0.19 | | 0.15 |
| **Neurologic/Vascular** | Peginterferon-lambda | 0.68 (0.47, 0.98) | 0.037 | 0.72 (0.53, 0.98) | 0.034 | 0.63 (0.48, 0.83) | 0.001 |
| | Placebo | 0.60 (0.36, 0.98) | 0.043 | 0.63 (0.43, 0.93) | 0.019 | 0.51 (0.34, 0.76) | 0.001 |
| | *Difference in decline* | | 0.52 | | 0.5 | | 0.32 |
| **Mood** | Peginterferon-lambda | 0.59 (0.29, 1.20) | 0.14 | 0.64 (0.35, 1.19) | 0.16 | 0.56 (0.30, 1.07) | 0.077 |
| | Placebo | 0.34 (0.12, 0.98) | 0.046 | 0.38 (0.13, 1.10) | 0.074 | 0.52 (0.29, 0.93) | 0.029 |
| | *Difference in decline* | | 0.14 | | 0.20 | | 0.88 |

**Table S3. The specific severe symptoms reported by participants in each treatment group.**

| PegIFN Lambda | ID 3 | ID 18 | ID 9 | ID 15 | ID 31 | ID 47 | ID 52 |
|---|---|---|---|---|---|---|---|
| **Day 0** | throat | | | smell | smell | smell | smell |
| **Day 0.5** | throat | | | | | smell | |
| **Day 1** | | | | | smell | smell | |
| **Day 2** | fever | | | | smell | smell | |
| **Day 3** | cough | | | | | smell | |
| **Day 4** | | | | | | | |
| **Day 5** | | | smell | | | | |
| **Day 6** | | cough, SOB | smell | | | | |
| **Day 7** | | | | | | | |
| **Day 10** | | | smell | | | | |
| **Day 12** | | | smell | | | | |
| **Day 14** | | cough, SOB | | | | | |

| Placebo | ID 5 | ID 11 | ID 16 | ID 27 | ID 33 | ID 51 | ID 60 |
|---|---|---|---|---|---|---|---|
| **Day 0** | chills | | fever, smell, nausea | fatigue | cough, fever, chills, nausea, vomit | smell | |
| **Day 0.5** | | | | | cough, throat | fatigue, headache | |
| **Day 1** | | | | | cough, fever, chills, throat, myalgia, abdo, vomit | smell | |
| **Day 2** | | | fatigue, nausea | | | fatigue, smell | |
| **Day 3** | | | fatigue, nausea | | | fatigue, smell | |
| **Day 4** | | fatigue | fatigue | | | | |
| **Day 5** | | fatigue | | | | | cough |
| **Day 6** | | | | | diarrhea | | |
| **Day 7** | | | | | | | |
| **Day 10** | | | | | | | |
| **Day 12** | | | | | | | |
| **Day 14** | | | | | | | |

Smell – loss of sense of smell/taste, SOB – shortness of breath; abdo – abdominal pain
