## Supplementary figures and images for "Peginterferon-lambda for the treatment of COVID-19 in outpatients"

### Supp Fig 1.tiff

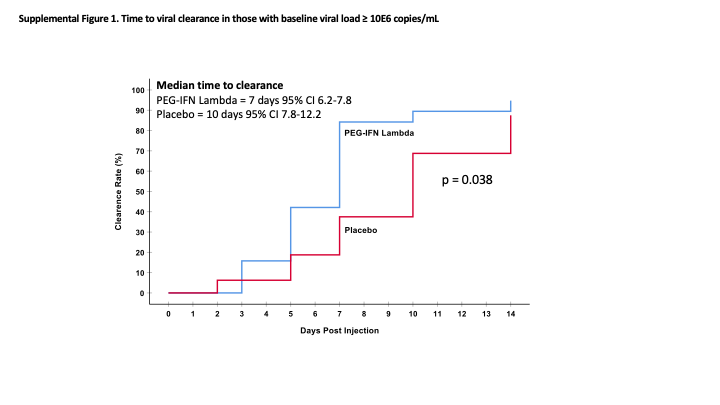

### Supp Fig 2.tiff

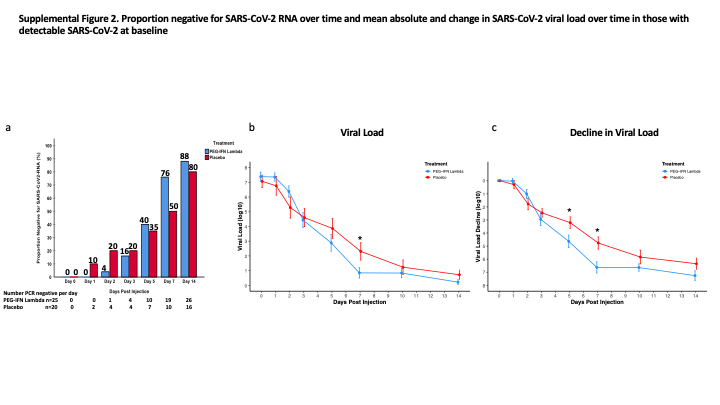

### Supp Fig 3.tiff

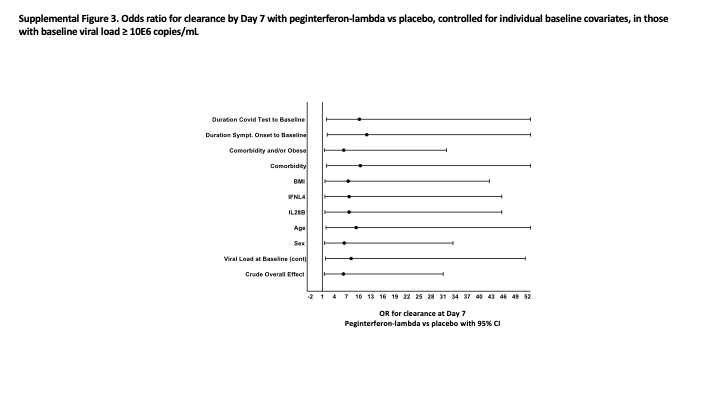

### Supp Fig 4.tiff

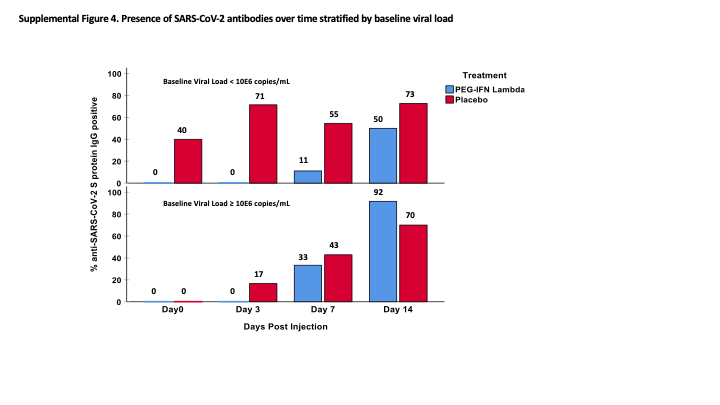

### Supp Fig 5.tiff

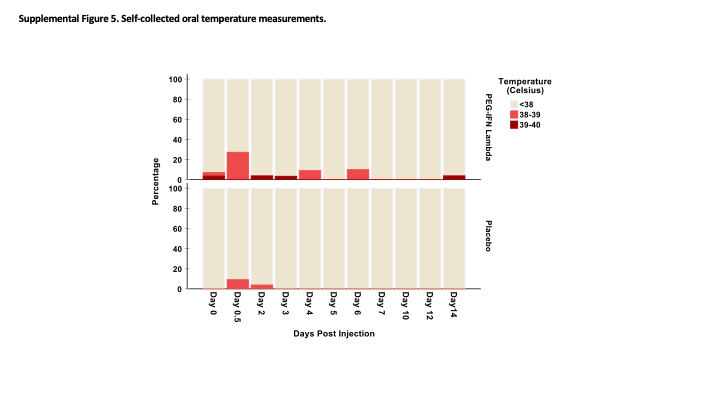
